# Dietary Macronutrient Intake and the Gut Microbiome in Adults Undergoing Bariatric Surgery for Obesity

**DOI:** 10.1101/2025.10.28.25338397

**Authors:** Sarah J. Lande, Lydia M. Stephney, Lucia Gonzalez A. Ramirez, Paula-Dene C. Nesbeth, Terryl J. Hartman, Dean P. Jones, Damaskini Valvi, Elizabeth M. Hechenbleikner, Edward Lin, Rob S. McConnell, Vaia Lida Chatzi, Jessica A. Alvarez, Thomas R. Ziegler

## Abstract

Limited information linking dietary intake to gut metagenomic data in bariatric surgery patients is available. We examined whether there were correlations between macronutrient intake and the gut microbiome and related gene pathways prior to and following bariatric surgery.

Participants were 29 adults living with obesity undergoing bariatric surgery (93% females). Three-day food records were analyzed prior to and after surgery to estimate mean daily intakes of macronutrients to derive measures of diet quality [glycemic index, added sugar intake, and the Healthy Eating Index 2015 (HEI-2015)]. Pre- and post-operative stool samples were sequenced using whole-genome shotgun sequencing to identify changes in microbial composition. Diversity indices and differential abundance were calculated, and correlations between dietary intake and outcomes were assessed using linear regression and machine learning models.

At the phylum level, pre-operative Synergistetes abundance was positively correlated with soluble fiber intake, and Proteobacteria was inversely linked with HEI-2015 scores. Post-operatively, Lentisphaerae was inversely correlated with dietary glycemic index. The change in Verrucomicrobia abundance was inversely correlated with the change in glycemic index, and the change in Fusobacteria abundance was positively correlated with the change in glycemic index. The changes in several functional gene pathways were positively linked to the change in HEI-2015 scores, the change in soluble fiber intake, and the change in insoluble fiber intake.

In adults undergoing bariatric surgery, intakes of specific macronutrients pre-operatively and as a function of the change after surgery were correlated with several microbial phyla, genera, and nutrient-related functional gene pathways.

---

Numerous studies have shown that a more diverse gut microbial composition is associated with better health outcomes [1]. The gut metagenome can be described based on microbiota composition (e.g., bacterial composition), species richness (number of different types of microorganisms present), species diversity (amount of each type of microbial species), and gene pathway enzyme abundance (e.g., amino acid metabolism pathways) [2].

Changes in dietary intake of protein, fat, carbohydrate (CHO), and other macronutrients, such as dietary fiber, can alter the gut microbiome in humans [3] [4]. For example, low-fiber diets, in comparison to high-fiber diets, are associated with higher levels of bile-tolerant microorganisms [3]. In older adults, higher carbohydrate quality (integration of the ratio of solid carbohydrates to total carbohydrates, dietary fiber, glycemic index, and the ratio of whole grains to total grains) was positively associated with Shannon alpha diversity [5]. Total energy, fat, and fiber intake also alter gut microbiome diversity and metabolism [6] [7].

Obesity itself is associated with dysbiosis of the gut microbiome and alterations in gut microbiota-associated functional gene pathways [8]. Bariatric surgery, including sleeve gastrectomy (SG) and Roux-en-Y gastric bypass (RYGB), is increasingly performed in adults living with obesity to mitigate or prevent morbidities, including type 2 diabetes and hypertension [9]. In one study, gut microbial richness was increased in the first year after bariatric surgery (RYGB or gastric banding) but tended to stabilize between one and five years after surgery [8]. Bariatric surgery has also been shown to alter functional gene pathways associated with the gut microbiota. After SG, metagenomic analysis showed greater ability to metabolize amino acids, while after RYGB, gut microbiota exhibited a greater ability for transport and metabolism of amino acids and carbohydrates [9,10]. Another study found that both RYGB and SG increased Proteobacteria, while Bacteroidetes increased in RYGB and decreased in SG six months after surgery [11].

Limited data have been published linking dietary intake to changes in the gut microbiome in bariatric surgery patients, and the published data are conflicting. In a Brazilian study, gut microbiota richness increased by three months after bariatric surgery and was positively correlated with fiber and inversely correlated with fat intakes, respectively [12]. In contrast, another study showed no significant relationship with dietary intake and microbiome indexes after bariatric surgery [13]. The purpose of this pilot study was to examine how macronutrient intake prior to and after bariatric surgery is linked to gut microbiome characteristics and nutrient-related functional gene pathways. We did so by examining the dietary intake of adults undergoing bariatric surgery and correlations to gut microbiome taxa and nutrient-related functional gene pathways using whole genome shotgun sequencing of stool samples.

## Materials and Methods

This study was conducted with a cohort of 29 adults living with obesity who were enrolled in a study to explore metabolic, body composition, and dietary endpoints before and after bariatric surgery, either RYGB or SG. Inclusion criteria were: 1) indication for elective RYGB or SG per National Institutes of Health (NIH) guidelines, namely body mass index (BMI) ≥ 40 kg/m^2^ or BMI ≥ 35 kg/m^2^ with at least one co-morbidity (hypertension, type 2 diabetes, dyslipidemia, or obstructive sleep apnea syndrome); 2) no prior gastric bypass surgery or gastrectomy; 3) age > 18 years. Exclusion criteria were: 1) evidence or history of acute or chronic infectious diseases (e.g., HIV, tuberculosis), inflammatory diseases (e.g., rheumatoid arthritis, inflammatory bowel disease), cancer (other than non-melanoma localized skin cancer), alcoholism; 2) history of severe psychosocial disorder within the previous year; 3) history of type 1 diabetes. This study was observational, not interventional; therefore, a Consolidated

Standards of Reporting Trials (CONSORT) diagram was not necessary. The study was approved by the Emory University Investigational Review Board (IRB00102564). Informed consent was obtained prior to bariatric surgery, and the study visits were conducted between 11/27/2018 and 11/17/2020. Participants were admitted to the Georgia Clinical and Translational Science Alliance (Georgia CTSA) Clinical Research Center (GCRC) at Emory University Hospital for a study visit within two months prior to the scheduled surgery and again six to ten months after bariatric surgery. Body weight was measured by trained research staff in the GCRC using a research-caliber scale, and height was measured without shoes using a research stadiometer to calculate BMI as kg/m^2^.

Due to the COVID-19 pandemic, several participants were unable to undergo their scheduled surgeries. Additionally, some follow-up evaluations post-operatively were unable to occur or were postponed past the scheduled date. These interruptions caused by the pandemic led to a smaller number of participants studied in the post-operative phase, or the post-operative visit occurring more than six to ten months after bariatric surgery.

The type of surgery was not considered in this study because the number of participants who received either RYGB or SG was too small to justify analyzing data by surgery type.

There was no adjustment for age because the ages of all participants were similar, and there was no adjustment for sex because there were only two males (**Table 1**).

**Table 1:** Demographic Data of Participants.

|  | **Pre-Operative**<br>**Cohort (n=29)** | **Pre- and Post-**<br>**Operative**<br>**Cohort (n=19)** |
| --- | --- | --- |
| Female, n (%) | 27 (93) | 17 (89) |
| Male, n (%) | 2 (7) | 2 (11) |
| Age, mean SD | 42.3 ± 11.4 | 42.9 ± 10.1 |
| Race, n (%) |  |  |
| Caucasian | 12 (41) | 8 (42) |
| AA/Black | 17 (59) | 11 (58) |
| Ethnicity, n (%) |  |  |
| Not Hispanic | 20 (69) | 13 (68) |
| Hispanic/Latino | 9 (31) | 6 (32) |
| Gastric sleeve operation, n (%) | N/A | 12 (63) |
| Roux-En-Y bypass operation, n (%) | N/A | 7 (36) |
| Baseline BMI, mean (kg/m<sup>2</sup>) ± SD | 49.4 ± 11.8 | 47.5 ± 7.7 |
| Change in BMI, mean kg/m<sup>2</sup> (%) |  | -9.9 (-20.9) |
| Prevalent Type 2 Diabetes Mellitus, n (%) | 1 (3) | 1 (5) |

AA= African American, BMI = body mass index, n=29 participants pre-operatively, n=19
participants pre- and post-operatively, SD = standard deviation

### Dietary Intake

Participants recorded their food and beverage intake over a span of three days (one weekend day and two weekdays) during a seven-day period prior to each GCRC outpatient admission. Each three-day food record was evaluated for completeness, and macronutrient intake was determined using Nutrition Data System for Research (NDS-R) software (University of Minnesota, Minneapolis, MN). The Healthy Eating Index 2015 (HEI-2015) was determined [14]. At the individual level, the intake of each macronutrient was divided by total calorie intake and then multiplied by 1000 as appropriate. The list of macronutrients analyzed in relation to metagenomic data is listed in **Supplemental Table 1**. It should be noted that participants were advised by their medical teams to be on a calorie-restricted diet for two weeks before bariatric surgery.

### Gut Microbiome Sequencing

Stool samples were collected the morning of each GCRC visit using the OMNIgene-GUT self-collection and stabilization system (DNAGenotek Inc., Ottawa, Ontario, Canada). Samples were sent to CosmosID (Germantown, MD), and DNA was extracted using the Qiagen DNeasy PowerSoil Pro kit according to the manufacturer’s protocol. Extracted samples were quantified using the Qubit 4 fluorometer and Qubit dsDNA HS Assay kit (Thermofisher Scientific). DNA libraries were prepared using the Nextera XT DNA Library Preparation Kit (Illumina) and IDT Unique Dual Indexes with total DNA input of 1ng. Genomic DNA was fragmented using a proportional amount of Illumina Nextera XT fragmentation enzyme. Unique dual indexes were added to each sample, followed by 12 cycles of PCR to construct libraries. DNA libraries were purified using AMpure magnetic Beads (Beckman Coulter) and eluted in QIAGEN EB buffer. DNA libraries were quantified using Qubit fluorometer and Qubit™ dsDNA HS Assay Kit. Whole-genome shotgun sequencing was performed on an Illumina NextSeq 2000 instrument using paired 250bp reads.

### Bioinformatics and Statistical Analyses

In this pilot study, we did not adjust results for age given the age range of participants or sex given that there were only two males (**Table** 1).

Dietary intake and microbiome/metagenomic data were analyzed to determine correlations between dietary intake of specific macronutrients and metagenomic data. Two-tailed paired t-tests were used to determine the differences in macronutrient intake between time points.

Sequencing data was analyzed using the CosmosID-HUB pipeline (detailed pipeline methods can be found in US patent US-10042976-B2). The system utilizes a high-performance data-mining k-mer algorithm that rapidly disambiguates millions of short sequence reads into the discrete genomes engendering the sequences. The pipeline has two separable comparators: the first consists of a pre-computation phase for reference databases, and the second is a per-sample computation. The inputs to the pre-computation phase are databases of reference genomes, virulence markers, and antimicrobial resistance markers that are continuously curated by CosmosID scientists. The output of the pre-computational phase is a phylogeny tree of microbes, together with sets of variable-length k-mer fingerprints (biomarkers) uniquely associated with distinct branches and leaves of the tree.

The second per-sample computational phase searches the hundreds of millions of short sequence reads, or contigs from draft *de novo* assemblies, against the fingerprint sets. This query enables the sensitive yet highly precise detection and taxonomic classification of microbial next-generation sequencing (NGS) reads. The resulting statistics are analyzed to return the fine-grain taxonomic and relative abundance estimates for the microbial NGS datasets. To exclude false positive identifications, the results are filtered using a filtering threshold derived based on internal statistical scores that are determined by analyzing many diverse metagenomes.

### Functional Analysis Methods

Initial quality control, adapter trimming, and preprocessing of metagenomic sequencing reads are done using BBduk [15]. The quality-controlled reads are then subjected to a translated search against a comprehensive and non-redundant protein sequence database,

UniRef90. The UniRef90 database, provided by UniProt [16], represents a clustering of all non-redundant protein sequences in UniProt, such that each sequence in a cluster aligns with 90% identity and 80% coverage of the longest sequence in the cluster. The mapping of metagenomic reads to gene sequences is weighted by mapping quality, coverage, and gene sequence length to estimate community-wide weighted gene family abundances as described by Franzosa et al [17]. Gene families are then annotated to MetaCyc [18] reactions (Metabolic Enzymes) to reconstruct and quantify MetaCyc metabolic pathways in the community as described by Franzosa et al. Furthermore, the UniRef90 gene families are also regrouped to GO terms [19] in order to get an overview of GO functions in the community. Lastly, to facilitate comparisons across multiple samples with different sequencing depths, the abundance values are normalized using total-sum scaling (TSS) normalization to produce “Copies per million” (analogous to TPMs in RNA-Seq) units.

### Statistical Analysis Methods

Filtered data for each sample is combined into a single feature table for analysis. This table is output from the CosmosID-HUB as abundance score data, which is analogous to count data. Abundance scores are imported into the statistical programming language R [20] (version 4.3.2) and normalized to relative abundance. The following statistical analyses and visualizations were performed:

### Figure Generation and Statistical Analysis

Heatmaps: Heatmaps were created using the pheatmap R package [21] generated using the phylum, genus, species, and strain matrices for bacteria from CosmosID-HUB. Heatmaps were created using the log scale ((−log(qvalue)*sign(beta coefficient)). Hierarchical clustering and dendrograms are generated using the hclust and dist functions from the base R stats package, with default parameters using Euclidean distance and complete linkage. Alpha Diversity Boxplots (with Wilcoxon Rank-Sum): Alpha diversity boxplots were calculated from the phylum, genus, species, and strain-level abundance score matrices from CosmosID-HUB analysis. Chao, Simpson, and Shannon alpha diversity metrics were calculated in R using the R package Vegan [22]. Wilcoxon Rank-Sum tests were performed between groups using the R package ggsignif [23]. Boxplots with overlaid significance in p-value format were generated using the R package ggpubr [24].

MaAsLin2: Microbiome Multivariable Associations with Linear Models (MaAsLin) [25] was implemented using the R package MaAsLin2. MaAsLin is designed to assess multivariable associations with microbiome community features with complex metadata. MaAsLin performs generalized linear and mixed models to accommodate a wide range of studies and data types (counts or relative abundance), including longitudinal and cross-sectional study designs. It was used to identify significant associations of metadata of interest with individual taxa. Fixed effects are accounted for in identifying significant associations with specific taxa and functions. False discovery rates are controlled for using the Benjamini-Hochberg procedure to adjust p-values for multiple comparisons (reported as q-values). MaAsLin also determined the beta coefficients for correlations between specific macronutrients and specific taxa and functional genes. Repeated measures are accounted for in the random effects term.

Boruta: Boruta [26] is a wrapper around a random forest machine learning algorithm. Boruta improves the baseline algorithm by calculating shadow variables from the data itself in order to determine whether variables are important in classifying a binary response. Boruta performs a top-down search for relevant features by comparing the importance of the original attributes to that of random permutations of the attributes. Irrelevant features were progressively eliminated to stabilize the test set.

ROC curves: Receiver operator characteristic (ROC) curves use test sensitivity and specificity to determine if a metric improves the classification of a binary response variable. The R package pROC [27] was used to plot the ROC curve. For both Boruta and ROC curves, data was split into test and train sets to optimize parameters using the training set before running the algorithm on the test set.

### Alpha Diversity

To analyze the impact of macronutrients on gut microbial alpha diversity indexes, we classified each participant’s intake as being above or below the median nutrient intake. Gut microbial alpha diversity was determined across sample groups at the species level using the CHAO1 and Shannon indexes [28]. Significance was determined using the Wilcoxon Rank Sum Test. The CHAO1 abundance index estimates the number of observed microbial species, and Shannon combines species richness and evenness, or the species number and abundance, respectively [28].

## Results

### Demographic Data

**Table 1** shows the demographic data of the participants. Most of the participants were female (93%), and the mean age (yrs) of the total of 29 participants was 42.3 ± 11.4. A total of 59% of participants were African American/Black, 41% were Caucasian, and 31% of the total participants identified as Hispanic/Latino. The mean time between surgery and the post-operative GCRC visit is 8.9 ± 2.8 months. The mean pre-operative BMI at the baseline visit was 49.4 ± 11.8 kg/m^2^. Participants lost approximately 21% of their body weight from the baseline visit to the post-bariatric surgery visit. The mean decrease in BMI was 9.9 kg/m^2^ from the baseline pre-operative to the post-operative study visit.

At the pre-operative visit, eight participants were taking oral proton pump inhibitors, and one was taking an oral antibiotic. At the post-operative visit, five participants were taking oral proton pump inhibitors, and three were taking oral antibiotics. Also, participants were advised not to take pre- or probiotics other than potential food sources of these for at least 30 days before study visit.

### Macronutrient Intake Data

**Table 2** shows the change in macronutrient intake and HEI-2015 scores before vs. after surgery (pre-operative data was subtracted from post-operative data). Mean energy intake decreased by approximately 19% in the post-operative period compared to the pre-operative period (p=0.002). Similarly, the percentage of calories consumed as carbohydrates and carbohydrates as g/1000 calories fell by nearly 13% and 49%, respectively (**Table 2**). The percent change in mean daily intakes of total fiber (g/1000 kcal) and insoluble dietary fiber (g/1000 kcal) significantly rose by approximately 19% and 22%, respectively. Percent change in the intake of saturated fatty acids (SFA), monounsaturated fatty acids (MUFA), and polyunsaturated fatty acids (PUFA), as percentages of total calories, each significantly increased (**Table 2**). Finally, the dietary glycemic index fell slightly but significantly, and the mean glycemic load decreased moderately (≈ 18% decline).

**Table 2:** Macronutrient Intake Pre-Operative, Post-Operative, and Change After Surgery.

| **Nutrient** | **Pre-Op ±**<br>**SD**<br>**(n=29)** | **Post-Op ±**<br>**SD**<br>**(n=19)** | **Change**<br>**(n=19)** | **Percent**<br>**Change**<br>**After**<br>**Surgery**<br>**(%)** | **P-value** |
| --- | --- | --- | --- | --- | --- |
| HEI-2015 Score | 56.4 ± 12.7 | 57.7 ± 12.9 | +1.3 | +2.3 | 0.19 |
| Energy (kcal) | 1688 ± 599 | 1362 ± 614 | -325.8 | -19.3 | 0.002 |
| % kcal as Fat | 38.5 ± 6.7 | 42.2 ± 8.8 | +3.7 | +9.6 | 0.049 |
| % kcal as CHO | 43.8 ± 7.9 | 38.2 ± 10.4 | -5.6 | -12.8 | 0.009 |
| % kcal as Protein | 18.1 ± 4.1 | 20.5 ± 5.3 | +2.4 | +13.3 | 0.03 |
| Fat (g)/1000 kcal | 42.8 ± 7.4 | 46.8 ± 9.8 | +4.0 | +9.3 | 0.048 |
| CHO (g)/1000 kcal | 186.3 ±<br>77.3 | 95.6 ± 25.9 | -90.7 | -48.7 | <0.001 |
| Pro (g)/1000 kcal | 45.3 ± 10.1 | 51.2 ± 13.2 | +5.9 | +13.0 | 0.03 |
| Total Fiber (g)/1000<br>kcal | 8.8 ± 3.2 | 10.5 ± 3.6 | +1.7 | +19.3 | 0.02 |
| Soluble Dietary Fiber<br>(g)/1000 kcal | 3.4 ± 2.3 | 3.8 ± 1.7 | +0.4 | +11.8 | 0.55 |
| Insoluble Dietary<br>Fiber (g)/1000 kcal | 5.4 ± 2.4 | 6.6 ± 2.8 | +1.2 | +22.2 | 0.02 |
| % Calories from SFA | 7.4 ± 3.5 | 9.6 ± 3.8 | +2.2 | +29.7 | 0.001 |
| % Calories from<br>MUFA | 9.4 ± 4.8 | 13.2 ± 5.0 | +3.8 | +40.4 | <0.001 |
| % Calories from<br>PUFA | 6.4 ± 3.1 | 8.7 ± 3.4 | +2.3 | +35.9 | <0.001 |
| Polyunsaturated to<br>Saturated Fat Ratio | 0.7 ± 0.5 | 0.9 ± 0.5 | +0.2 | +28.6 | 0.25 |
| Omega-3 Fatty Acids<br>(g)/1000 kcal | 1.2 ± 0.5 | 1.3 ± 0.6 | +0.1 | +8.3 | 0.20 |
| Omega-6 Fatty Acids<br>(g)/1000 kcal | 9.2 ± 2.6 | 10.0 ± 2.7 | +0.8 | +8.7 | 0.24 |
| Glycemic Index<br>(glucose reference) | 61.2 ± 5.2 | 58.2 ± 5.8 | -3.0 | -4.9 | 0.004 |
| Glycemic Load<br>(glucose<br>reference)/1000 kcal | 61.5 ± 15.8 | 50.2 ± 16.9 | -11.3 | -18.4 | 0.001 |
| Added Sugars (by<br>Total Sugars)<br>(g)/1000 kcal | 23.7 ± 13.1 | 17.0 ± 12.7 | -6.7 | -28.3 | 0.03 |
| Whole Grains (ounce<br>equivalents)/1000 kcal | 0.4 ± 0.3 | 0.5 ± 0.4 | +0.1 | 25.0 | 0.17 |
| Refined Grains<br>(ounce<br>equivalents)/1000 kcal | 2.6 ± 1.1 | 2.2 ± 1.2 | -0.4 | -15.4 | 0.10 |

Note: A total of 19 participants had both pre-operative and post-operative dietary intake data.
CHO= carbohydrate, MUFA = monounsaturated fatty acids, Pro = protein; PUFA=
polyunsaturated fatty acids; SFA = saturated fatty acids

### Alpha Diversity Data

#### Pre-Operative

Gut microbial alpha diversity in relation to nutrient intake showed that the median intake of both PUFA (% calories) and the polyunsaturated to saturated fat ratio were significantly and positively correlated with gut microbial alpha diversity using the CHAO1 index (**Table 3**).

**Table 3:**
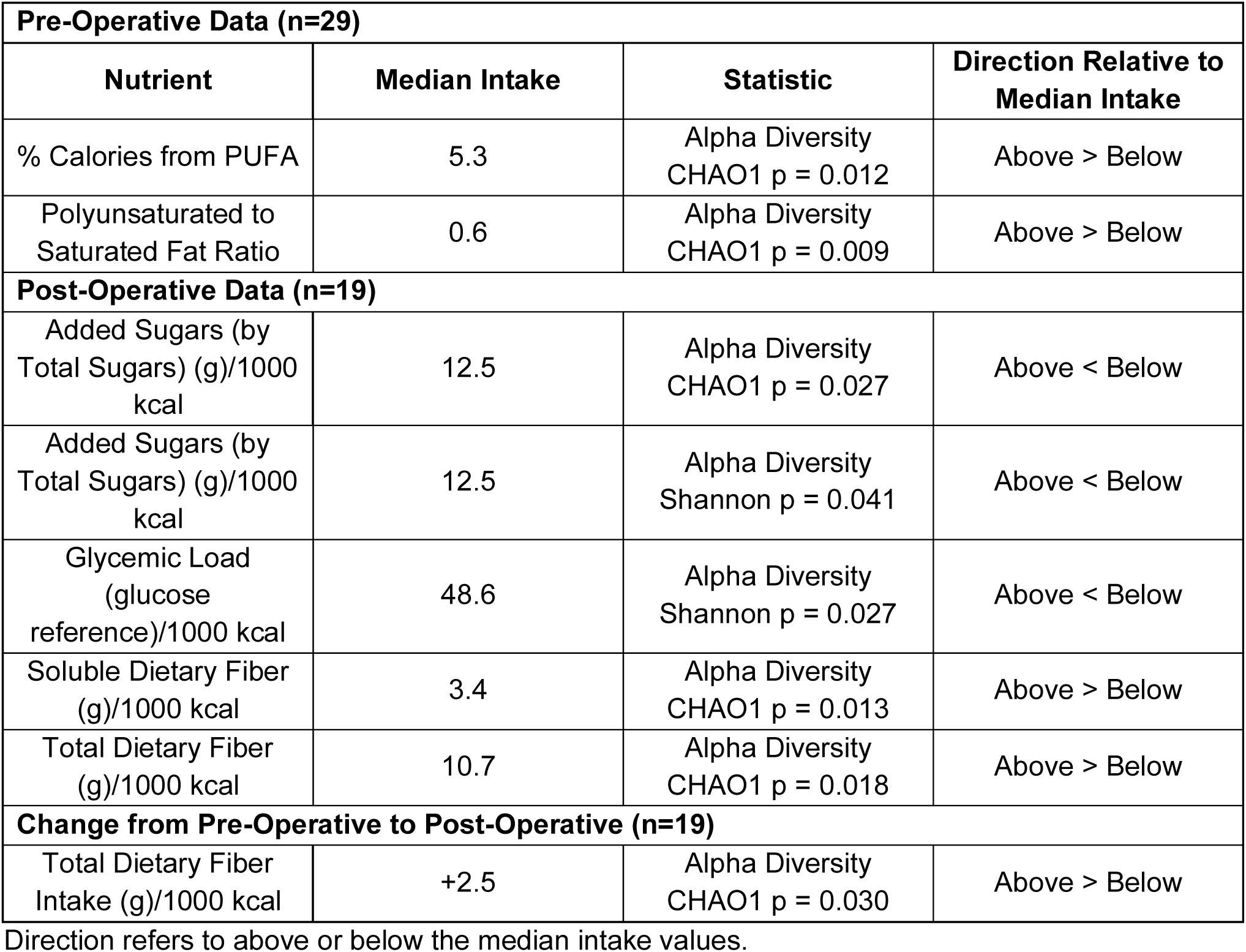
Microbial Alpha Diversity in Relation to Median Nutrient Intake.

#### Post-Operative

Gut microbial alpha diversity in relation to median nutrient intake in the post-operative period showed that the median intake of added sugar (g/1000 kcal) was inversely correlated with alpha diversity using the CHAO1 and Shannon indexes (**Table 3 and Figure 1**). Also, dietary glycemic load was inversely correlated with alpha diversity by the Shannon index. In contrast, median intake of soluble fiber (g/1000 kcal) and total fiber (g/1000 kcal) were significantly and positively correlated with alpha diversity using the CHAO1 index.

**Figure 1.**
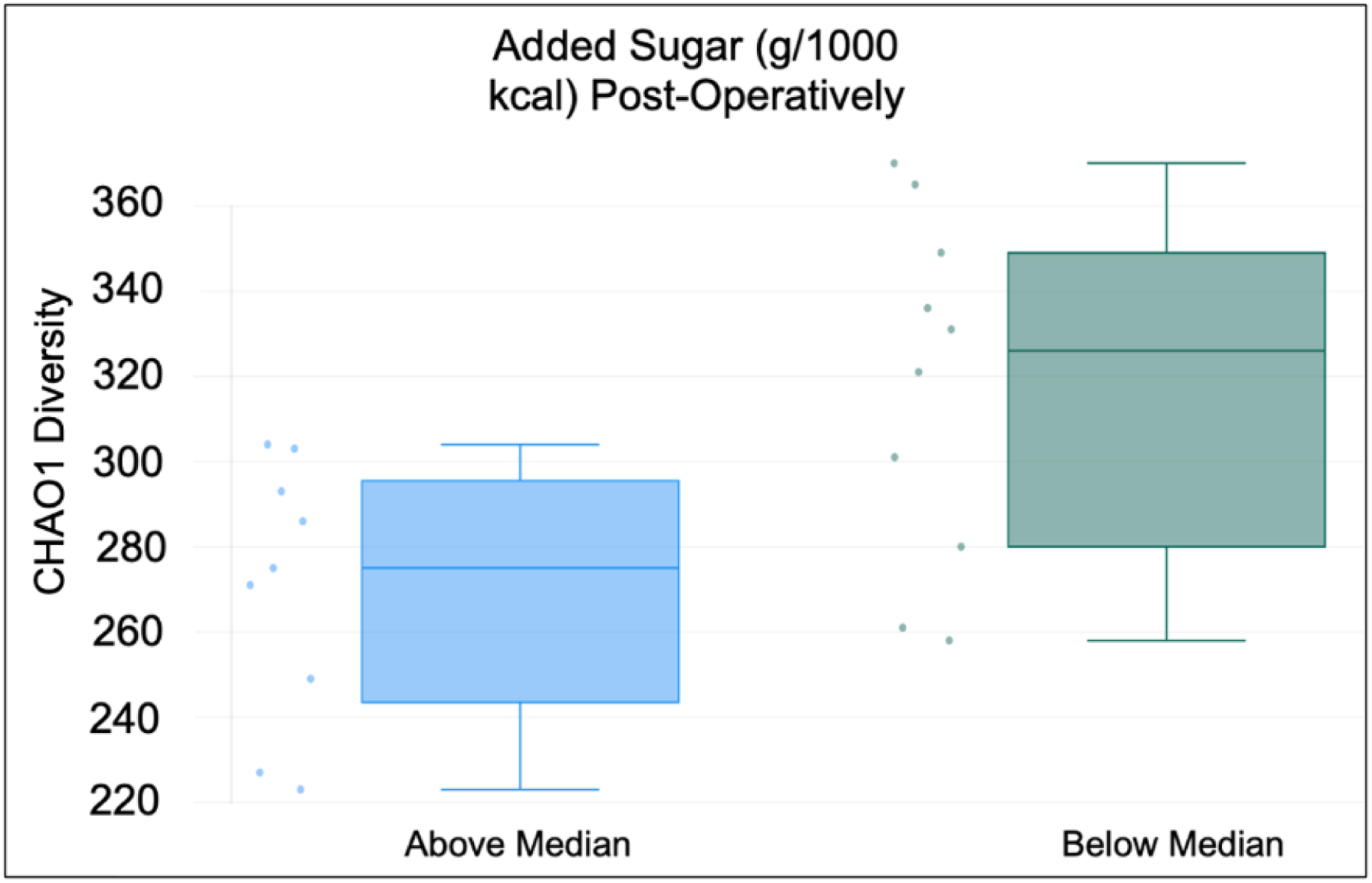
Comparison of alpha diversity based on median added sugar intake. Dietary intake data was collected using three-day food records and NDS-R software. Alpha-diversity was determined by the CHAO1 diversity index. Results are from 19 participants in the post-operative phase after bariatric surgery.

#### Change as a Function of Surgery

The change in total dietary fiber (g/1000 kcal) was positively correlated with alpha diversity using the CHAO1 index (**Table 3**).

### Taxa Abundance Data

#### Pre-Operative

**Table 4** shows macronutrient intakes and HEI-2015 scores in the pre-operative (n=29) period in relation to the taxa data (**Table 4**). At the phylum level, we found that Synergistetes was positively correlated with soluble dietary fiber intake (g/1000kcal), and polyunsaturated to saturated fat ratio. Actinobacteria was positively correlated with MUFA, as a percentage of total calories. Proteobacteria was inversely correlated with HEI-2015 scores.

**Table 4:**
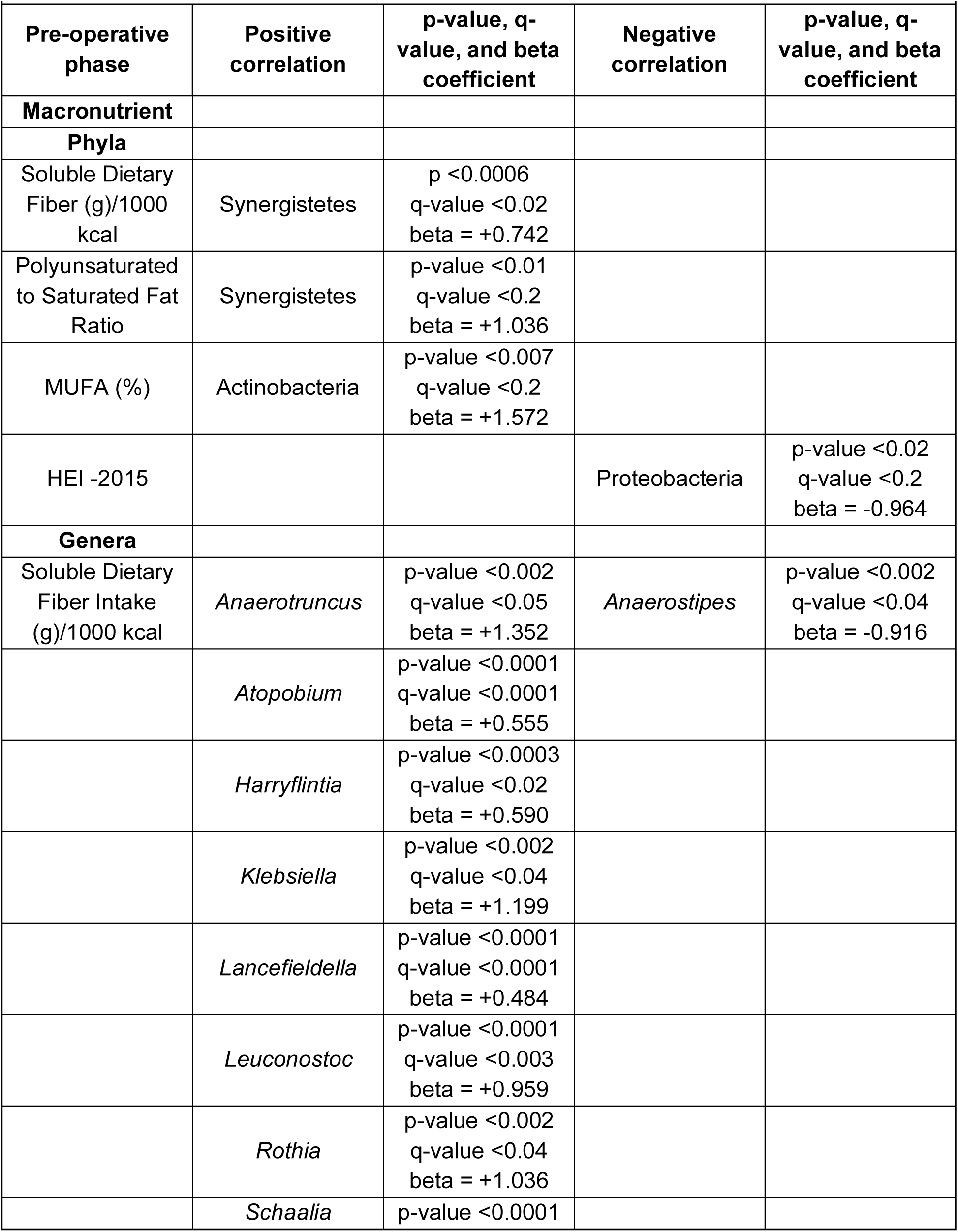
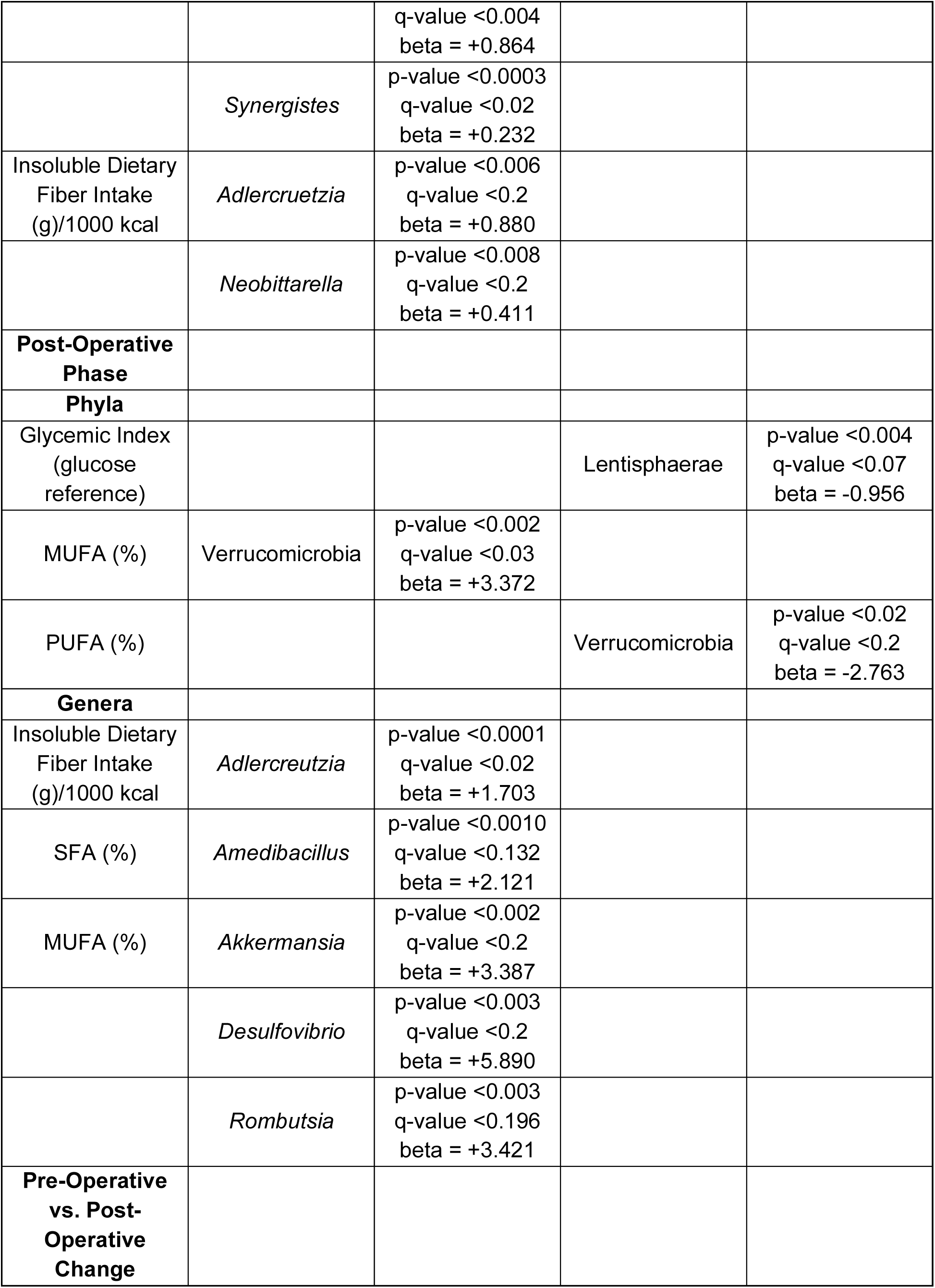
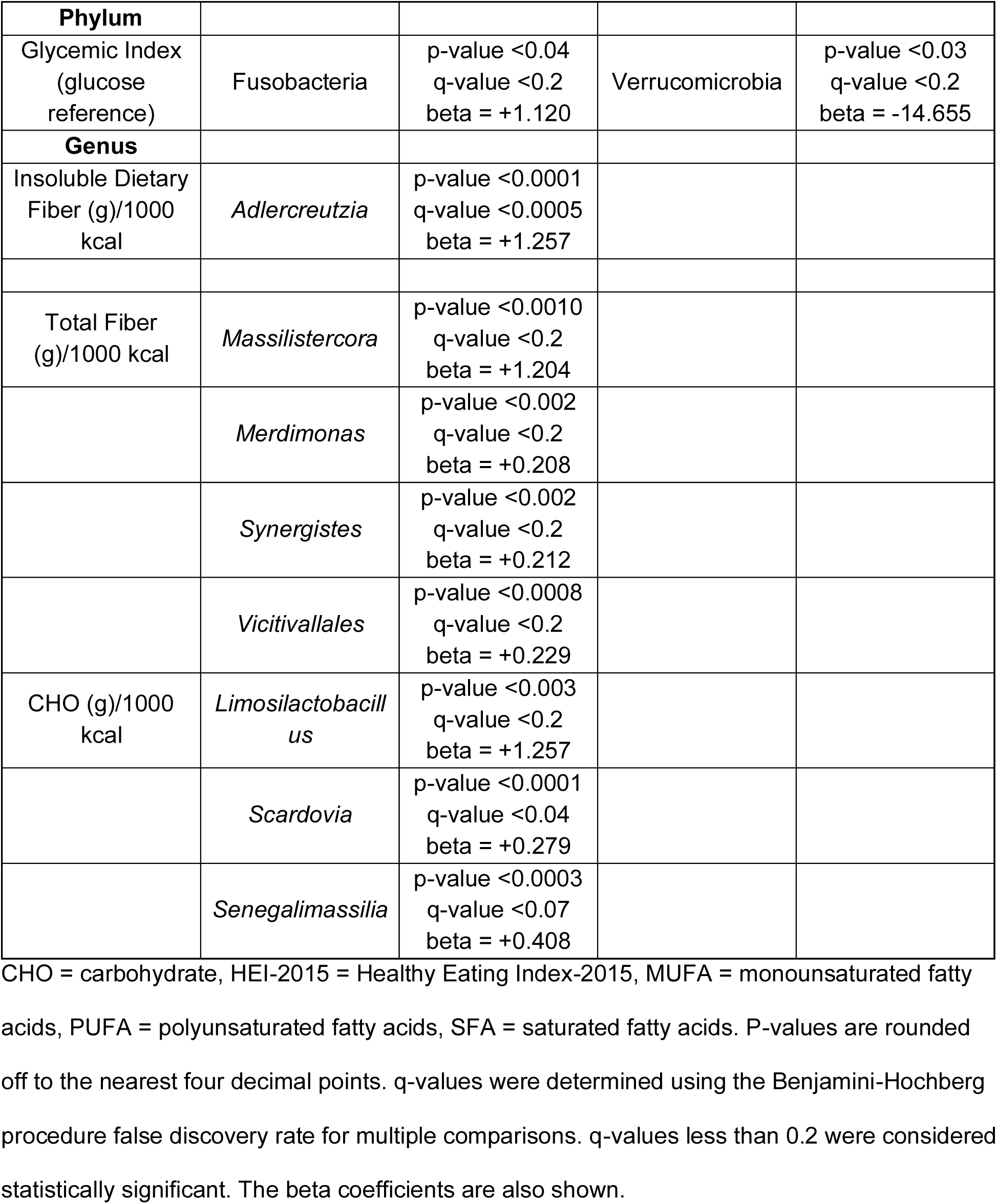
Dietary Macronutrient Intake and Gut Microbiota Correlations.

At the genus level, *Anaerotruncus, Atopobium, Harryflintia, Klebsiella, Lancefieldella, Leuconostoc, Rothia, Schaalia,* and *Synergistes* were positively correlated with soluble dietary fiber intake (g/1000 kcal). *Anaerostipes* was inversely correlated with soluble dietary fiber intake (g/1000 kcal). *Adlercruetzia* and *Neobittarella* were positively correlated with insoluble dietary fiber intake (g/1000 kcal).

#### Post-Operative

At the phylum level, post-operative Lentisphaerae was inversely correlated with glycemic index score. Verrucomicrobia was positively correlated with MUFA as a percentage of total calories and inversely correlated with PUFA as a percentage of total calories.

At the genus level, *Adlercreutzia* was positively correlated with insoluble dietary fiber intake (g/1000 kcal). *Amedibacillus* was positively correlated with SFA as a percentage of total calories. *Akkermansia, Desulfovibrio,* and *Rombutsia* were positively correlated with MUFA as a percentage of total calories.

#### Change as a Function of Surgery

We next evaluated the change in macronutrient intake and HEI-2015 scores from pre-operative to post-operative (n=19) in relation to the change in taxa. At the phylum level, the change in Verrucomicrobia was inversely correlated, and the change in Fusobacteria was positively correlated with the change in glycemic index score (**Figure 2**).

**Figure 2.**
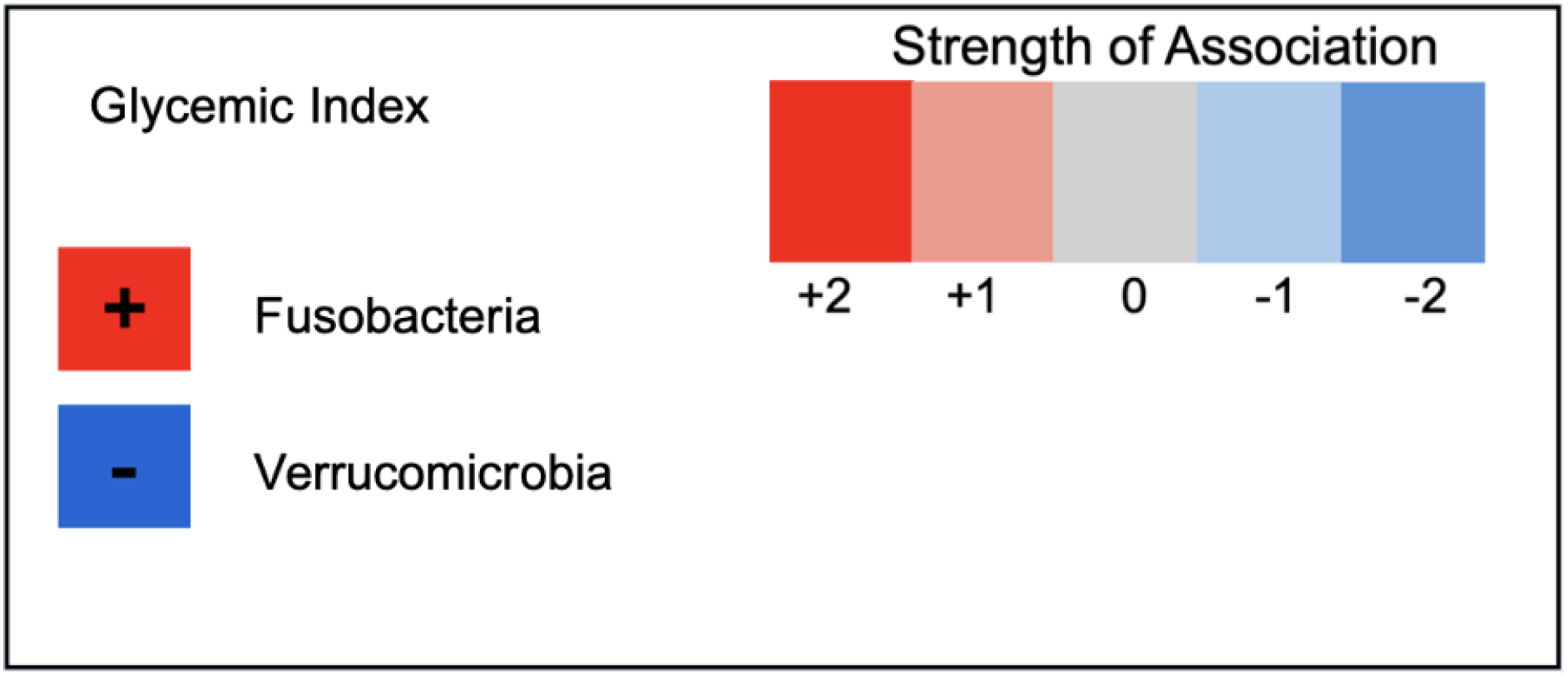
Heatmap of change in the phyla Fusobacteria and Verrucomicrobia correlated with change in glycemic index pre- and post-surgery. Dietary intake data was collected using three-day food records, and glycemic index was determined using NDS-R software. The data were correlated with gut microbiome taxa in 19 participants undergoing bariatric surgery. The strength of the correlations is shown in the color-coded heatmap.

At the genus level, the change in *Adlercreutzia* was positively correlated with the change in insoluble dietary fiber (g/1000kcal) intake. The changes in *Massilistercora, Merdimonas, Synergistes,* and *Vicitivallales* were positively correlated with the change in total fiber (g/1000 kcal) intake. The changes in *Limosilactobacillus, Scardovia,* and *Senegalimassilia* were positively correlated with the change in CHO (g/1000 kcal) intake.

### Gene Pathway Abundance Data

#### Pre-Operative

Nutrient-related functional pathways using GO and MetaCyc were analyzed to determine correlations with macronutrient intake. Soluble dietary fiber (g/1000 kcal) was positively correlated with copper ion binding, superpathway of L-arginine and L-ornithine degradation, and superpathway of L-threonine metabolism. Soluble dietary fiber intake (g/1000 kcal) was inversely correlated with several pathways, including glucose catabolic process, galactoside-2-alpha L-fucosyltransferase, and the thiamine biosynthetic process. Insoluble dietary fiber intake (g/1000 kcal) was inversely correlated with ferroxidase activity and positively correlated with S-adenosyl L-methionine cycle I (**Table 5**).

**Table 5:**
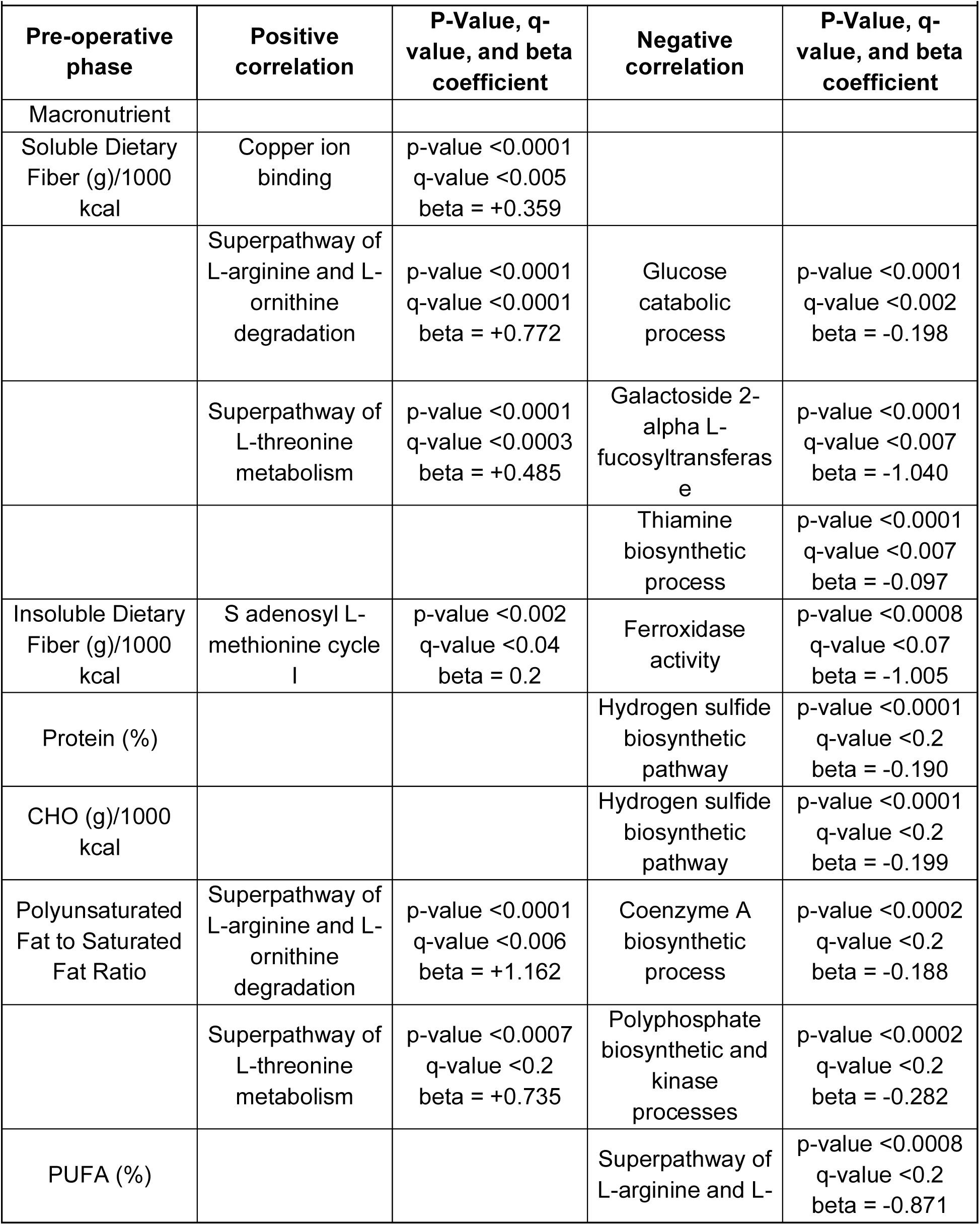
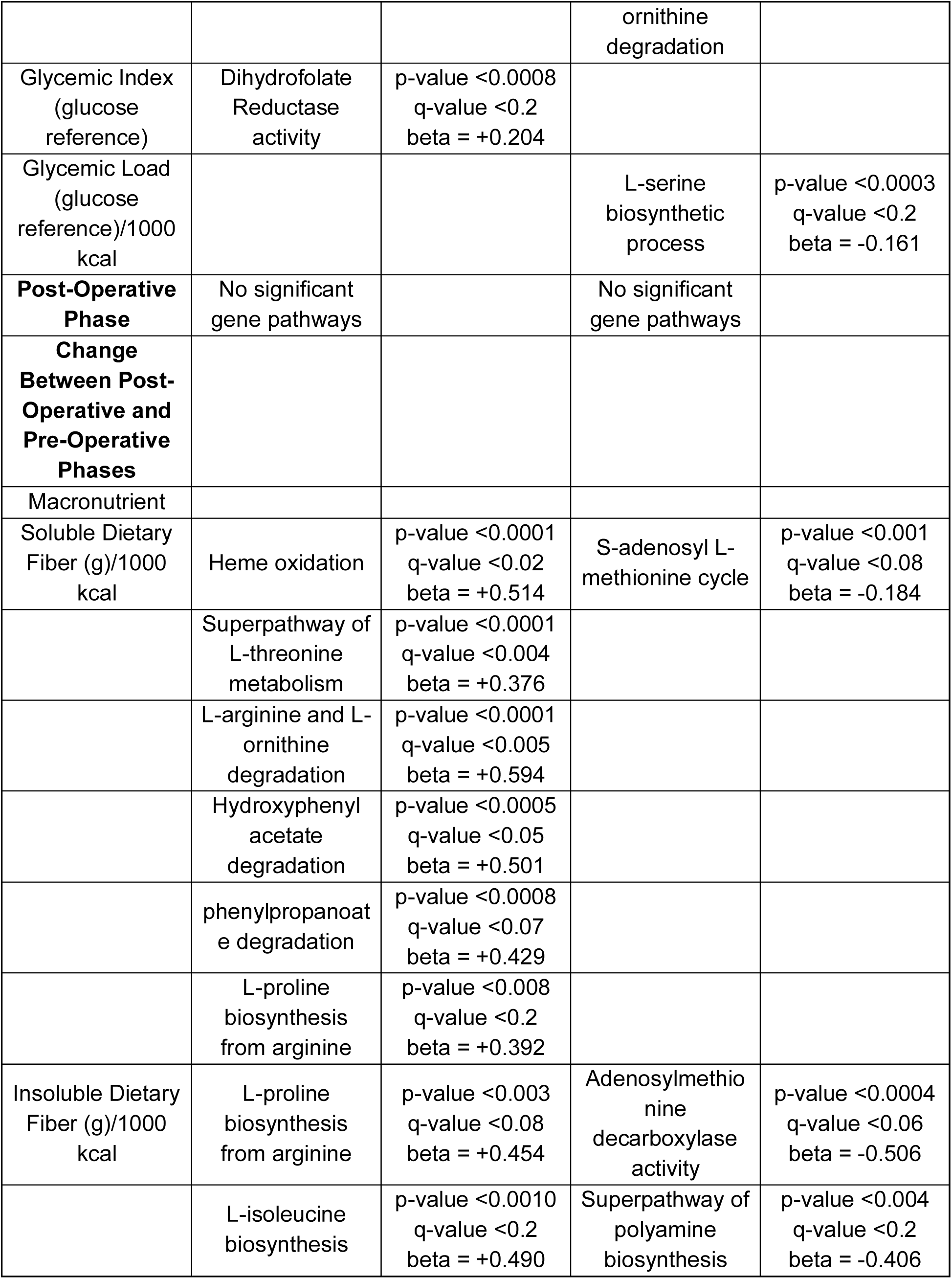
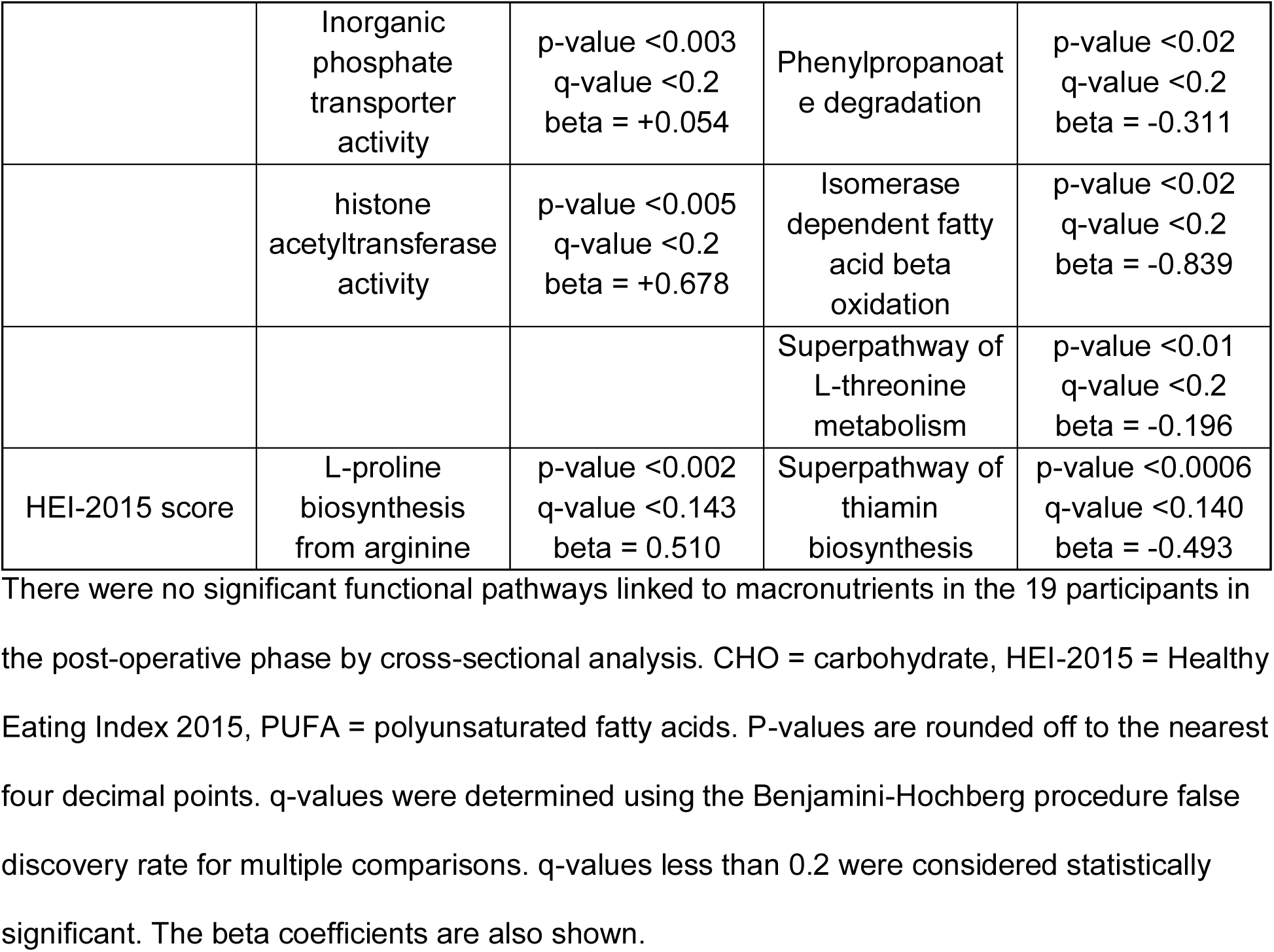
Nutrient-Related Functional Genes and Correlations with Macronutrient Intake.

Protein intake as a percentage of total calories and CHO intake (g/1000 kcal) were each inversely correlated with the hydrogen sulfide biosynthetic pathway. The following functional pathways were inversely correlated with the polyunsaturated fat to saturated fat ratio: coenzyme A biosynthetic process and polyphosphate biosynthetic, and kinase processes. The polyunsaturated fat to saturated fat intake ratio was positively correlated with the superpathway of L-arginine and L-ornithine degradation, and the superpathway of L-threonine metabolism.

PUFA intake as a percentage of total calories was inversely correlated with the superpathway of L-arginine and L-ornithine degradation. Glycemic index score was positively correlated with dihydrofolate reductase activity, and glycemic load was inversely correlated with L-serine biosynthetic process (**Table 5**).

#### Post-Operative

No significant relationships with macronutrient intakes and nutrient-related gene pathways were observed.

#### Change as a Function of Surgery

The change in soluble dietary fiber intake (g/1000 kcal) was positively correlated with the changes in heme oxidation, superpathway of L-threonine metabolism, L-arginine and L-ornithine degradation, hydroxyphenylacetate, phenylpropanoate degradation, and L-proline biosynthesis from arginine (**Figure 3**). Conversely, the change in soluble dietary fiber intake (g/1000 kcal) was inversely correlated with the change in S-adenosyl L-methionine cycle. The change in insoluble dietary fiber intake (g/1000 kcal) was positively correlated with the changes in L-proline biosynthesis from arginine (**Figures 3 and 4**), L-isoleucine biosynthesis, and inorganic phosphate transporter activity. The change in insoluble dietary fiber intake (g/1000 kcal) was inversely correlated with the changes in adenosylmethionine decarboxylase activity, superpathway of polyamine biosynthesis, phenylpropanoate degradation, isomerase-dependent fatty acid beta oxidation, superpathway of L-threonine metabolism, and histone acetyltransferase activity (**Table 5**).

**Figure 3.**
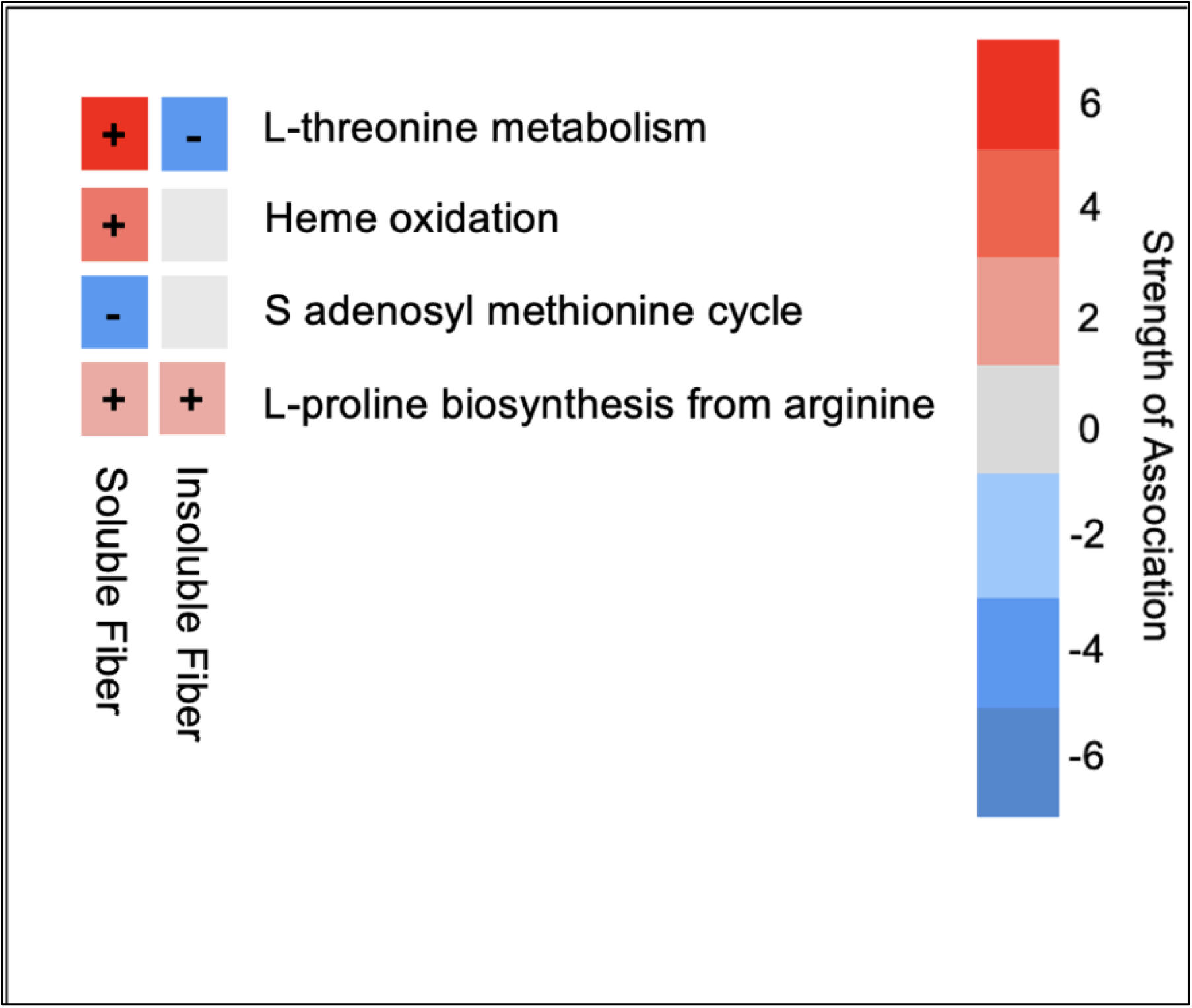
Heatmap of change in selected nutrient-related pathways correlated with change in soluble and insoluble fiber intake pre- and post-surgery. Dietary intake data was determined using three-day food records and NDS-R software. Nutrient-related gene pathways were determined in 19 participants undergoing bariatric surgery. The strength of the correlations is shown in the color-coded heatmap.

**Figure 4.**
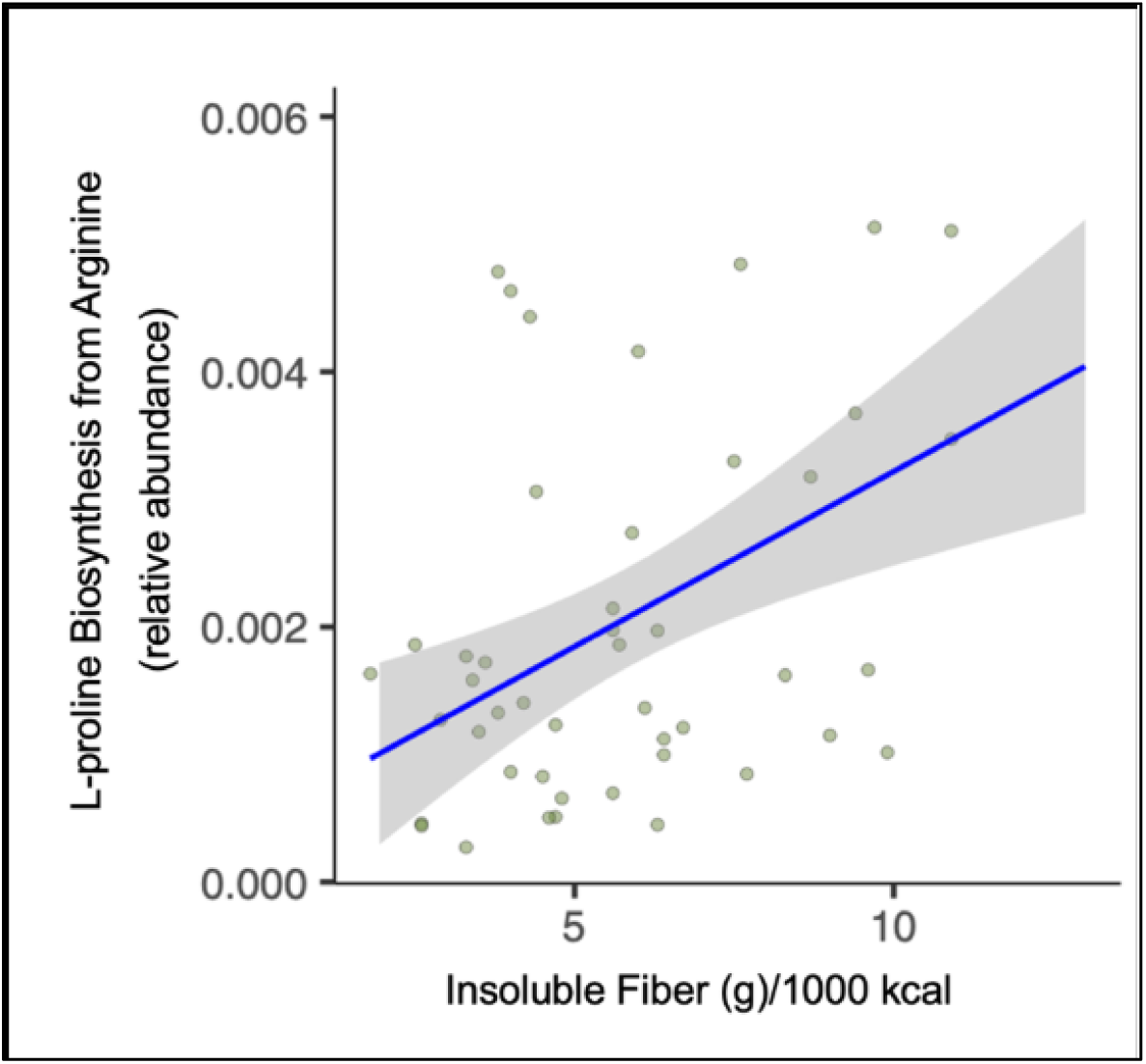
Relationship between change in insoluble fiber intake and change in L-proline biosynthesis from arginine as a function of surgery. Dietary intake data was collected using three-day food records and determined using NDS-R software. Nutrient-related gene pathways were determined by stool metagenomic analysis in 19 participants undergoing bariatric surgery. Change was determined as a difference in values from pre- to post-bariatric surgery.

The change in HEI-2015 scores was positively correlated with the change in L-proline biosynthesis from arginine and inversely correlated with the change in thiamin biosynthesis.

### Taxa Changes as a Function of Surgery

#### Phyla

At the phylum level, the change in CHO intake (% of total kcal) and the change in CHO intake (g/1000 kcal) were significant predictors of the change in pre-operative and post- operative gut microbiome profiles **(Figure 5A)**. We performed an ROC curve analysis to determine the validity of these predictions. This revealed an area under the curve (AUC) of 93.1% (**Figure 5B**) for both CHO intake as a percent of total calorie intake and CHO intake in grams/1000 calories. This indicates excellent predictive power for the change in these CHO intake categories (**Supplemental Table 1**), predicting changes in the overall gut microbiome profile as a function of surgery.

**Figure 5.**
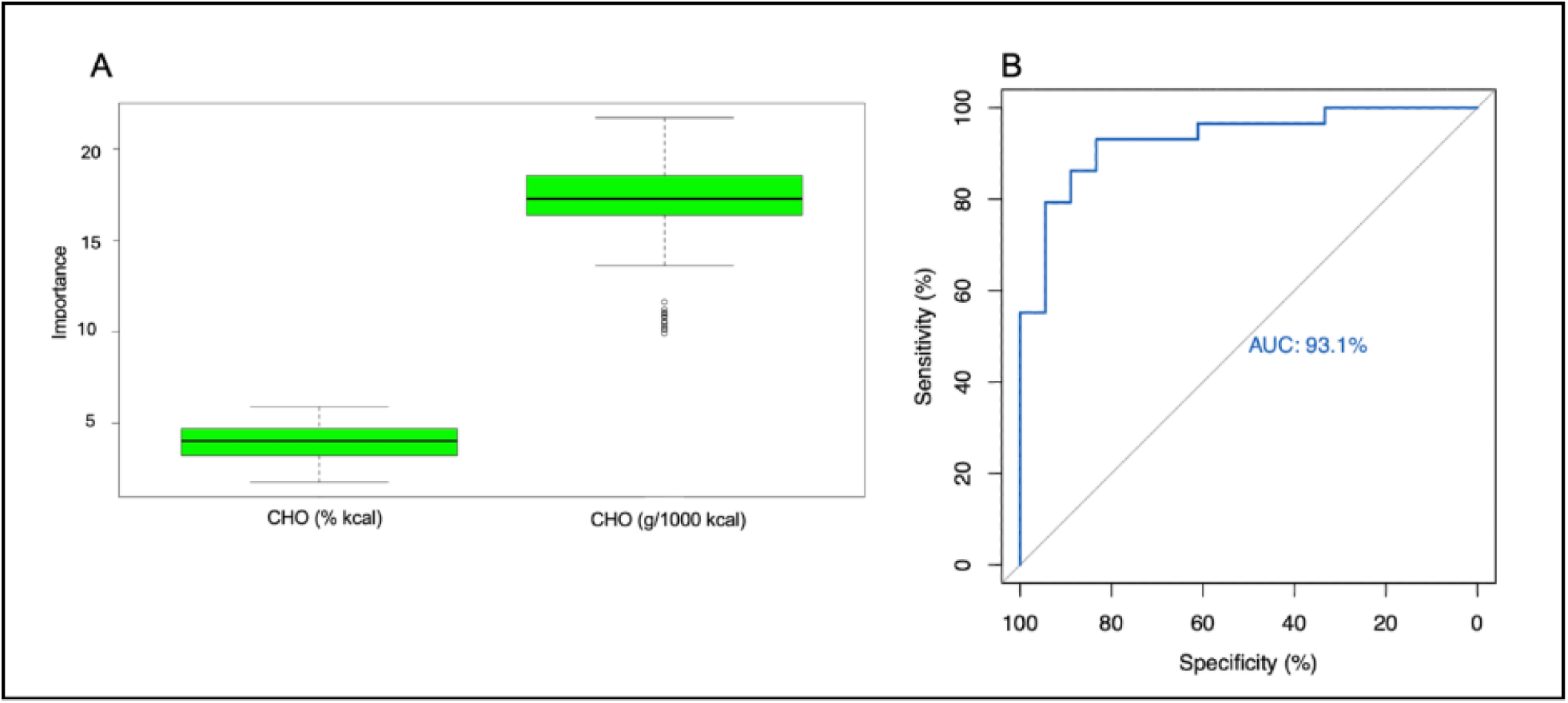
Identification of features that were significant predictors of change in the overall gut microbiome profile and evaluation of those predictions at the phylum level. a. CHO (% kcal) and CHO (g/1000 kcal) intake were significant predictors of change in pre- operative vs. post-operative gut microbiome profiles at the phylum level. Dietary intake data was collected using three-day food records and determined using NDS-R software. Dietary macronutrients and phyla were analyzed to determine which factors were predictors of change in the gut microbiome of 19 participants undergoing bariatric surgery. b. Receiver operating characteristic (ROC) curve evaluating the validity of the predictors of change in the gut microbiome profile at the phylum level. The outputs of the Boruta algorithm were evaluated to determine if they were adequate predictors of change in the gut microbiome profile of 19 participants undergoing bariatric surgery.

#### Genera

At the genus level, the change in CHO (g/1000 kcal) intake and the genus *Rothia* were predictors of pre-operative and post-operative differences in the microbiome profile **(Figure 6A)**. These predictions were confirmed with an ROC curve with an AUC of 80.5% **(Figure 6B)**, which indicates excellent predictive power and confirms that *Rothia* and CHO (g/1000 kcal) are predictors of change in the overall microbiome profile before and after surgery.

**Figure 6.**
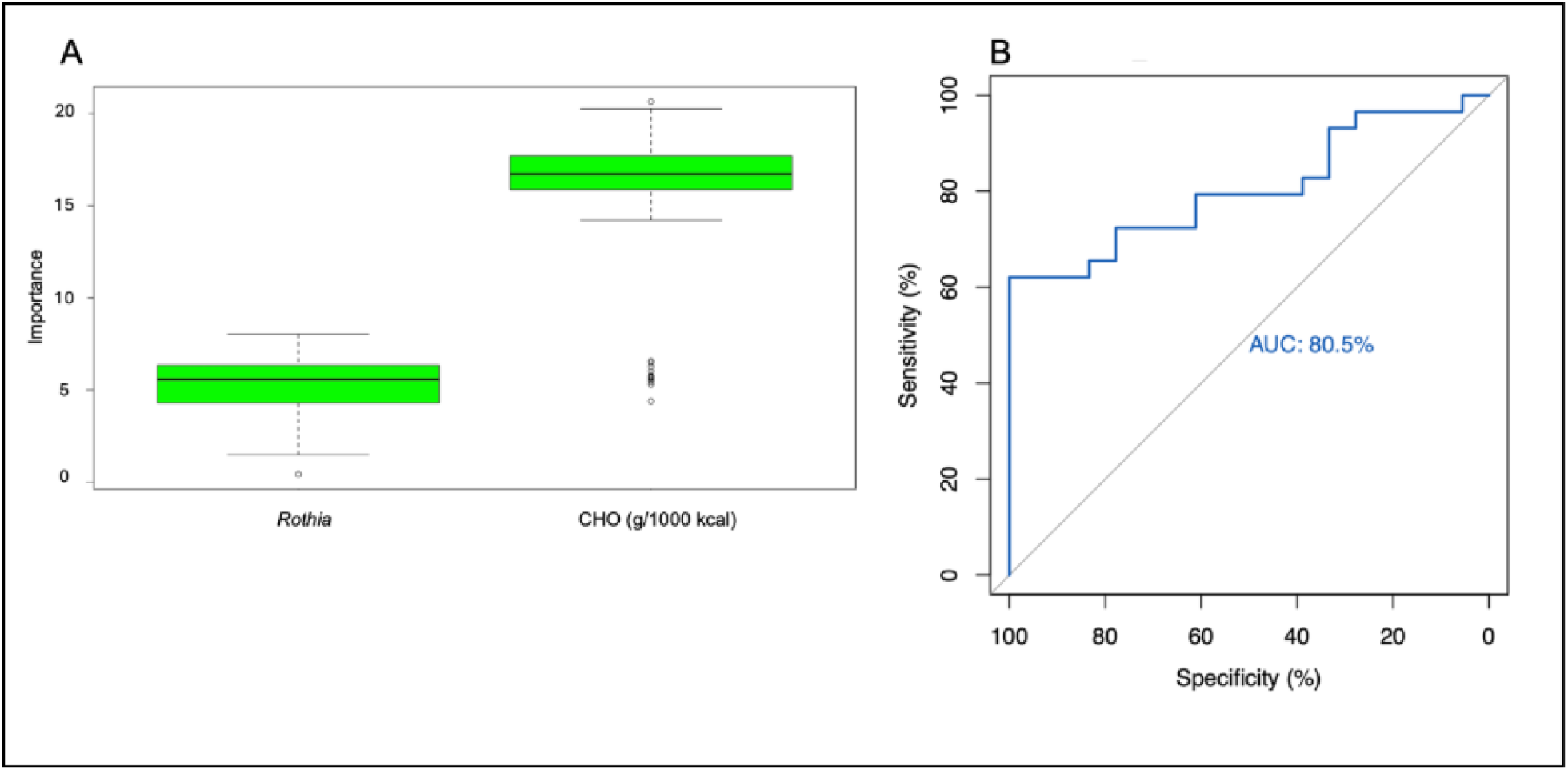
Identification of features that were significant predictors of change in the overall gut microbiome profile and evaluation of those predictions at the genus level. a. *Rothia* and CHO (g/1000 kcal) intake were significant predictors of change in pre-operative vs. post-operative microbiome profiles at the genus level. Dietary intake data was collected using three-day food records and determined using NDS-R software. Dietary macronutrients and genera were analyzed to determine which factors were predictors of change in the gut microbiome of 19 participants undergoing bariatric surgery. b. Receiver operating characteristic (ROC) curve evaluates the validity of the predictors of change in the gut microbiome profile at the genus level. The outputs of the Boruta algorithm were evaluated to determine if they were adequate predictors of change in the gut microbiome profile of 19 participants undergoing bariatric surgery.

### Nutrient-Related Functional Gene Pathway Changes as a Function of Surgery

CHO (g/1000 kcal) and four functional pathways (superpathway of purine *de novo* biosynthesis, S-adenosyl L-methionine cycle, cellulase activity, and dipeptidase activity) were significant predictors of pre-operative to post-operative differences in the overall gut microbiome profile.

The predictions were confirmed by an 87.5% to 92.3% AUC predicted from an ROC curve, indicating excellent predictive power.

## Discussion

In this pilot study, several microbial phyla, genera, and nutrient-related gene pathways were correlated with dietary intake of specific macronutrients and indexes of carbohydrate quality in the pre-operative and post-operative phases of bariatric surgery. Intakes of specific lipids, added sugars, glycemic load, and soluble and total dietary fiber were correlated with alpha diversity indexes. Indexes of dietary fiber intake, followed by indexes of fat intake, and then carbohydrate intake, were all correlated with various taxa **(Table 4)**. Both soluble and insoluble fiber intakes were the macronutrient class correlated with the most nutrient-related functional gene pathways **(Table 5)**. Change in total carbohydrate intake was the macronutrient class (as a percentage of total calories and in g/1000 kcal) identified as a significant predictor of change in the overall microbial composition. Numerous nutrient-related functional genes were correlated with macronutrient intake in the pre-operative phase and with changes between the pre- and post-operative phases.

In individuals living with obesity and not undergoing bariatric surgery, gut microbial alpha diversity is lower compared to normal-weight individuals [29]. We found that added sugar intake was inversely correlated with alpha diversity post-operatively. This is consistent with data showing that high simple sugar intake correlates with decreased gut microbial diversity [30].

Also, excess dietary sugar intake has been linked to gut microbial dysbiosis [31]. It has been documented that high dietary sugar intake is associated with decreased gut microbial diversity, given that excess monosaccharides are metabolized by specific taxa that favor simple carbohydrates [32]. The increase in such bacteria with higher added sugar intakes would serve to decrease alpha diversity.

We found total and soluble dietary fiber intake to be positively correlated with alpha diversity **(Table 3)**, consistent with data of Assal et al., in which dietary fiber was correlated with increased gut microbial richness in bariatric surgery patients in the post-operative vs. the pre- operative period [12]. Dietary fiber intake is associated with increased gut microbial alpha diversity, likely due to fermentation by numerous bacterial species [33] [34] [35]. We also found that soluble and insoluble fiber intakes were correlated with changes in the largest number of nutrient-related functional gene pathways (**Table 5**).

There was a positive association with the phylum Synergistetes and intake of soluble fiber and polyunsaturated to saturated fat ratio in the pre-operative phase. Synergistetes comprises several genera and species of gram-negative, anaerobic bacteria [36], which have been associated with type 2 diabetes, obesity, and other inflammatory conditions [37] [38] [38]. Although increased soluble fiber intake and consumption of a diet with a higher polyunsaturated to saturated fatty acid ratio are considered healthy dietary practices, these data may be counterintuitive. However, to our knowledge, there is no previous data linking Synergistetes with macronutrient intake.

The phylum Actinobacteria was positively correlated with dietary MUFA intake in the pre- operative phase. In a study comparing the gut microbiome of obese versus lean twins (77 dyads), Turnbaugh et al., found that 75% of obesity-related genes were derived from Actinobacteria [1]. Our findings are consistent with previous studies reporting that the abundance of Actinobacteria increased when participants were fed a MUFA-rich diet [39].

Higher Proteobacteria abundance was correlated with lower diet quality as determined by the HEI-2015 score in the pre-operative phase. In a long-term study of a general population cohort from California and Hawaii, a significant inverse association of Proteobacteria and HEI-2015 scores was found, in agreement with our findings [40]. Proteobacteria has been associated with obesity and type 2 diabetes, and an increase in Proteobacteria may be linked to gut dysbiosis [41]. Additionally, dysbiosis characterized by the change in Proteobacteria has been associated with inflammatory bowel disease [42].

Numerous genera were positively linked with pre-operative soluble dietary fiber intake **(Table 4)**. For example, *Anaerotruncus* was positively correlated with soluble dietary fiber intake. In healthy adults, a high saturated fat/low fiber diet was correlated with an increase in *Anaerotruncus*, a short-chain fatty acid producing genus associated with obesity [43]. Also, *Atopobium* was increased in association with increased soluble dietary fiber intake. This genus is more abundant in healthy individuals compared to those with type 2 diabetes and is inversely linked with BMI and inflammatory indices [44]. Species of the genus *Klebsiella*, such as *Klebsiella pneumoniae,* are associated with gut dysbiosis and disorders such as inflammatory bowel disease [45]. We found that soluble dietary fiber intake was positively correlated with *Klebsiella* in our participants with obesity as their only known inflammatory condition.

*Anaerostipes* was inversely linked with pre-operative soluble dietary fiber intake **(Table 4)**. *Anaerostipes* metabolizes luminal substrates such as soluble fiber to the short-chain fatty acid butyrate, which has cytoprotective properties and is an energy source for gut epithelia [46] [47].

We found that the post-operative dietary glycemic index was correlated with a higher abundance of bacteria in the phylum Lentisphaerae **(Table 4)**, a phylum which is of low abundance in people living with obesity [48–50].

Verrucomicrobia abundance is decreased in obese individuals [51]. Verrucomicrobia was positively correlated with MUFA and inversely correlated with PUFA dietary intake in the post-operative phase after weight loss induced by bariatric surgery. In our post-operative cohort, dietary MUFA intake increased by approximately 40% from the pre-operative period and was positively correlated with the genus *Akkermansia*. *Akkermansia* is a genus in the Verrucomicrobia phylum, and its abundance is increased in lean individuals and in people living with obesity while they are dieting [52]. Also, in a study of dietary caloric restriction, the species *Akkermansia muciniphilia* increased in association with weight loss [53] [54].

Gut Fusobacteria is associated with obesity [55] [56]. The decrease in glycemic index after bariatric surgery **(Table 2)** was correlated with a decrease in gut Fusobacteria abundance **(Table 3)**. The participants’ BMI decreased markedly by an average of 9.9 kg/m^2^ (−20.9%) after bariatric surgery (**Table 1**). To our knowledge, this is the first observation linking dietary glycemic index with this major obesity-associated phylum.

Regarding nutrient-related functional genes, in the pre-operative phase, soluble dietary fiber intake was positively correlated with copper ion binding, several superpathways of amino acid metabolism, and inversely correlated with genes related to thiamine and carbohydrate metabolism (**Table 5**). In addition, several other nutrient-related gene pathways were variously related to pre-operative intake of insoluble dietary fiber, protein, PUFA/SFA ratio, PUFA, glycemic index, and glycemic load.

The change in soluble dietary fiber intake (which increased post-operatively) was linked to regulation of several amino acid gene pathways, and upregulation of phenylpropanoate degradation, suggesting a link between microbial-derived short-chain fatty acid metabolism (**Table 5**).

Strengths of this study include new data linking dietary intake and gut metagenomic variables in obese individuals undergoing bariatric surgery. State-of-the-art dietary intake and metagenomic analysis were used. Limitations are the small number of participants (only one of whom had type 2 diabetes), the cross-sectional nature of the pre-operative analysis, and the pragmatic nature of the study, which included individuals undergoing either SG or RYGB surgeries. The study as designed is not generalizable to non-bariatric surgery patients.

## Conclusions

We conclude that in adults living with obesity undergoing bariatric surgery, specific macronutrient intake and HEI-2015 diet quality scores were correlated with several microbial taxa and nutrient-related functional gene pathways. Further work in larger prospective cohorts is needed to define specific regulated microbial species within the altered phyla and genera as a function of macronutrient intake, bariatric surgery type, and clinical characteristics.

## Data Availability

All data produced in the present study are available upon reasonable request to the authors

## Acknowledgments

We acknowledge the research contributions of the nurses and bionutrition staff of the Georgia Clinical and Translational Science Alliance Clinical Research Center (GCRC) at Emory University Hospital. We also thank Dr. Dana Walsh of CosmosID Inc. for her helpful contributions.

## Abbreviations

CHO: Carbohydrate
CONSORT: Consolidated Standards of Reporting Trials
Georgia CTSA: Georgia Clinical and Translational Science Alliance
GCRC: Georgia Clinical Research Center
HEI-2015: Healthy Eating Index 2015
MaAsLin: Microbiome Multivariable Associations with Linear Models
MUFA: Monounsaturated fatty acids
NIH: National Institutes of Health
NGS: Next-generation sequencing
NDSR: Nutrition Data System for Research
PUFA: Polyunsaturated fatty acids
ROC: Receiver operator characteristic
RYGB: Roux-en-Y gastric bypass
SFA: Saturated fatty acids
SG: Sleeve gastrectomy
TSS: Total-sum scaling

---

**Supplemental Table 1:** Mean Daily Dietary Intake of Macronutrients and Dietary Quality Determined Before and After Bariatric Surgery.

|  |  |
| --- | --- |
| Overall Dietary Quality | Healthy Eating Index (HEI) 2015 |
| Caloric Intake | Energy (kcal) |
| Indexes of Fat Intake | % Kcal as Fat (%) |
|  | Fat (g)/1000 kcal |
|  | % Calories from MUFA (%) |
|  | % Calories from PUFA (%) |
|  | % Calories from SFA (%) |
|  | Polyunsaturated to Saturated Fat Ratio |
|  | Omega-3 Fatty Acids (g)/1000 kcal |
|  | Omega-6 Fatty Acids (g)/1000 kcal |
| Indexes of Protein Intake | % Kcal as Protein (%) |
|  | Protein (g)/1000 kcal |
| Indexes of Carbohydrate Intake | % Kcal as Carbohydrate (%) |
|  | Carbohydrate (g)/1000 kcal |
|  | Glycemic Index (glucose reference) |
|  | Glycemic Load (glucose reference)/1000 kcal |
|  | Added Sugars (by Total Sugars) (g)/1000 kcal |
| Indexes of Grain Intake | Whole Grains (ounce equivalents)/1000 kcal |
|  | Refined Grains (ounce equivalents)/1000 kcal |
| Indexes of Fiber Intake | Total Fiber (g)/1000 kcal |
|  | Soluble Dietary Fiber (g)/1000 kcal |
|  | Insoluble Dietary Fiber (g)/1000 kcal |

MUFA = monounsaturated fatty acids; PUFA = polyunsaturated fatty acids; SFA = saturated fatty acids

